# STrain Analysis and Mapping of the Plantar Surface (STAMPS) – A novel technique of plantar load analysis during gait

**DOI:** 10.1101/2023.03.10.23287086

**Authors:** Alexander D. Jones, Sarah R. Crossland, Jane E. Nixon, Heidi J. Siddle, David A. Russell, Peter R. Culmer

## Abstract

Diabetic foot ulceration is driven by peripheral neuropathy, resulting in abnormal foot biomechanics and elevated plantar load. Plantar load comprises normal pressure and tangential shear stress. Currently, there are no in-shoe devices measuring both components of plantar load. The STAMPS (STrain Analysis and Mapping of the Plantar Surface) system was developed to address this and utilises digital image correlation (DIC) to determine the strain sustained by a plastically deformable insole, providing an assessment of plantar load at the foot-surface interface during gait.

STAMPS was developed as a multi-layered insole, comprising a deformable mid-layer, onto which a stochastic speckle pattern film is applied. A custom-built imaging platform is used to obtain high resolution pre- and post-walking images. Images are imported into commercially available DIC software (GOM Correlate, 2020) to obtain pointwise strain data. The strain and displacement data are exported and post-processed with custom analysis routines (MATLAB, Mathworks Inc.), to obtain the resultant global and regional peak strain (S_MAG_), antero-posterior strain (S_AP_) and medio-lateral strain (S_ML_). To validate the core technique an experimental test process used a Universal Mechanical Tester (UMT) system (UMT TriboLab, Bruker) to apply controlled vertical and tangential load regimes to the proposed multi-layer insole. A pilot study was then conducted to assess the efficacy of using the STAMPS system to measure in-shoe plantar strain in three healthy participants. Each participant walked 10 steps on the STAMPS insole using a standardised shoe. They also walked 10 m in the same shoe using a plantar pressure measurement insole (Novel Pedar®) to record peak plantar pressure (PPP) as a gold-standard comparator.

The results of the experimental validation tests show that with increased normal force, at a constant shear distance, S_MAG_ increased in a linear fashion. Furthermore, they showed that with increased shear distance, at a constant force, S_MAG_ increased. The results of the pilot study found participant 1 demonstrated greatest S_MAG_ in the region toes 3-5 (15.31%). The highest mean S_MAG_ for participant 2 was at the hallux (29.31%). Participant 3 exhibited highest strain in the regions of the 1^st^ and 2^nd^ metatarsal heads (58.85% and 41.62% respectively). Increased PPP was strongly associated with increased S_MAG_ with a Spearman’s correlation coefficient 0.673 (p <0.0001).

This study has demonstrated the efficacy of a novel method to assess plantar load across the plantar surface of the foot. Experimental testing validated the sensitivity of the method to both normal pressure and tangential shear stress. This technique was successfully incorporated into the STAMPS insole to reliably measure and quantify the cumulative degree of strain sustained by a plastically deformable insole during a period of gait, which can be used to infer plantar loading patterns. Future work will explore how these measures relate to different pathologies, such as regions at risk of diabetic foot ulceration.

## Introduction

Diabetic foot disease is a major global health concern. Over four hundred and sixty three million people live with diabetes worldwide,^1^ up to one quarter will develop a diabetic foot ulcer (DFU) and 5-8% will require a major amputation within one year.^2^ Development of a DFU is a multifactorial process driven by diabetic peripheral neuropathy (DPN) and peripheral arterial disease. DPN causes sensory, motor and autonomic dysfunction. Motor dysfunction manifests as muscle weakness leading to structural abnormalities in the foot, including hammer toe, claw toe and hallux valgus resulting in elevated tissue stress.^3^ In the presence of sensory neuropathy, this elevated tissue stress remains undetected, leading to persistent inflammation and eventual tissue breakdown.^4^

Plantar load comprises vertical and tangential components.^5^ The vertical component, plantar pressure, is well described in the literature. Patients with DPN exhibit elevated plantar pressure, with values commonly exceeding 600kPa, compared with 400 – 500kPa in those without foot pathology.^6,7^ Utilising pressure assessment to guide offloading strategies has been shown to reduce DFU healing time and recurrence,^8^ and is recommended by the International Working Group for the Diabetic Foot (IWGDF) to guide offloading strategies.^9^ Despite these guidelines, plantar pressure assessment is rarely performed outside of the research environment. This is in part due to the cost of the sensing devices and due to the time and expertise required to use them. Furthermore, whilst it has been demonstrated that those who develop DFUs often sustain elevated levels of plantar pressure, many do not, and there is often disparity between location of peak pressure and ulcer formation.^10^ Shear stress forms the tangential component of plantar load.^11^ Friction between the foot-surface interface causes shear stress in both antero-posterior and medio-lateral axes acting perpendicular to the long axis of the foot.^11^ Plantar shear stress was first described by Pollard et al. in 1983,^12^ however due to the difficulty in its measurement, remains poorly understood.^13^

The development of reliable technology to measure both plantar pressure and plantar shear stress has remained elusive, and no commercial platforms or wearable systems are available. Some centres have developed custom-made research platforms. The Cleveland clinic shear plate utilises an array of 80 tri-axial strain gauge sensors and has been used to measure shear and pressure in patients with diabetes.^14^ However, as a sensing platform it is limited to barefoot measurements which do not reflect the typical stresses experienced in-shoe. A recent systematic review conducted by the authors investigating plantar shear stress assessment found 16 studies investigating shear stress in patients with diabetes,^13^ with only three studies performing in-shoe analysis. Amemiya et al., used individual tri-axial piezoelectric sensors affixed to the sole of the foot.^15^ However, the sensor’s size prohibited assessment of the whole plantar surface, and as the sensors were not embedded within the insole they may act as foreign bodies. Lord et al., embedded three resistive sensors within an insole, though this again lacked the capacity to measure pressure and shear throughout the plantar surface.^16^ We have previously presented evidence-based requirements for wearable systems to monitor plantar load in patients with diabetic foot disease.^17^ Describing load measuring capabilities of >740kPa for pressure and >140 kPa for shear, distribution of sensors across the plantar surface, a sensor maximum surface area of 10mm x 10mm and a sampling rate of no less than 50Hz. In addition, sensors should be low profile and robust to maintain structural integrity in an environment subjected to significant load, changes in pH and temperature.^17^ To date, no systems approaching these requirements have been developed.

In this paper we propose a novel solution to address the need for a plantar load device. The principle of our approach is to use a plastically deformable insole to capture the cumulative level of strain at the foot-surface interface and to quantify this using Digital Image Correlation (DIC) to track strain before and after a period of gait. DIC is an optical based technique utilised to measure the resultant displacement, strain and deformation of materials after exposure to different load regimes.^18^ More commonly associated with material science, civil and aerospace engineering, DIC is increasingly being used with soft materials in the field of clinical biomechanics.^19^ Studies have successfully demonstrated its application investigating strain of aortic tissue, cartilage and sclera.^20–22^ It has recently been used to analyse strain on the plantar surface of the foot and found principle strain to be between 10-20% at the metatarsal heads during stance phase.^23^ The process of DIC entails comparing an image of the object of interest prior to loading, with those after or during loading.^18^ Image analysis algorithms then identify congruent blocks of pixels between the images to measure displacement or strain. If the structure lacks an inherent pattern to track, a stochastic speckle pattern is applied to the surface. DIC has several advantages over traditional sensing techniques. It is convenient and robust as it exploits conventional digital photography technology and does not require low vibration environments.^24^ It also has the capability to deliver high spatial resolution^14^ defined by the size and spacing of the speckle pattern and camera resolution.

In this work we present the STrain Analysis and Mapping of the Plantar Surface (STAMPS) system, an approach which harnesses DIC to measure and quantify the cumulative degree of shear strain sustained by a deformable insole during a period of gait, using this as the basis to infer plantar loading patterns. In *System Development* we present development of the STAMPS approach, *Experimental Evaluation of STAMPS* then details experimental validation prior to a reported *Pilot Study of STAMPS* with healthy participants to show the efficacy of the technique. We conclude with a discussion considering relevant limitations, future development and application of STAMPS in the clinical domain.

## System Development

### Requirements and Concept

The aim of the STAMPS system is to achieve an insole that deforms plastically when subjected to representative plantar loading regimes and that this deformation can be reliably tracked using DIC. The insole is designed to be a single-use measurement device due to the nature of the permanent plastic deformation it incurs during use. Furthermore, the STAMPS insole should be capable of covering the full plantar surface across a range of shoe sizes and shapes, whilst remaining sufficiently ‘low profile’ that it avoids disrupting natural gait patterns and causing dorsal complications.

To meet these aims, the overall concept of STAMPS is to fabricate a custom shoe insole and combine with an imaging platform, DIC workflow and post-hoc analysis routines, to determine personalised metrics on plantar strain. The STAMPs insole forms the core of this concept and comprises a multi-layered sheet, as shown in Figure 1, which can be cut to a desired size and shape. The mid-layer provides the deformable element of the insole upon which is a thin speckle-patterned film, to provide a trackable surface for DIC. A base layer acts as a supporting ‘scaffold’ to prevent distortion of the insole during fitting and retrieval. After pre-imaging, the insole is placed into a shoe and used over a series of gait cycles after which it is removed and processed with DIC to extract a strain map across the surface. In this context, the strain is a product of both plantar pressure and plantar shear stress. Thus, the level of strain, as measured by DIC, is the cumulative effect of plantar pressure and shear stress during the period of walking. Post-hoc analysis can then segment data into anatomical regions and create summary metrics for comparative studies. The following subsections detail how each aspect of the STAMPS concept was developed to realise the implementation for practical use.

**Figure 1:**
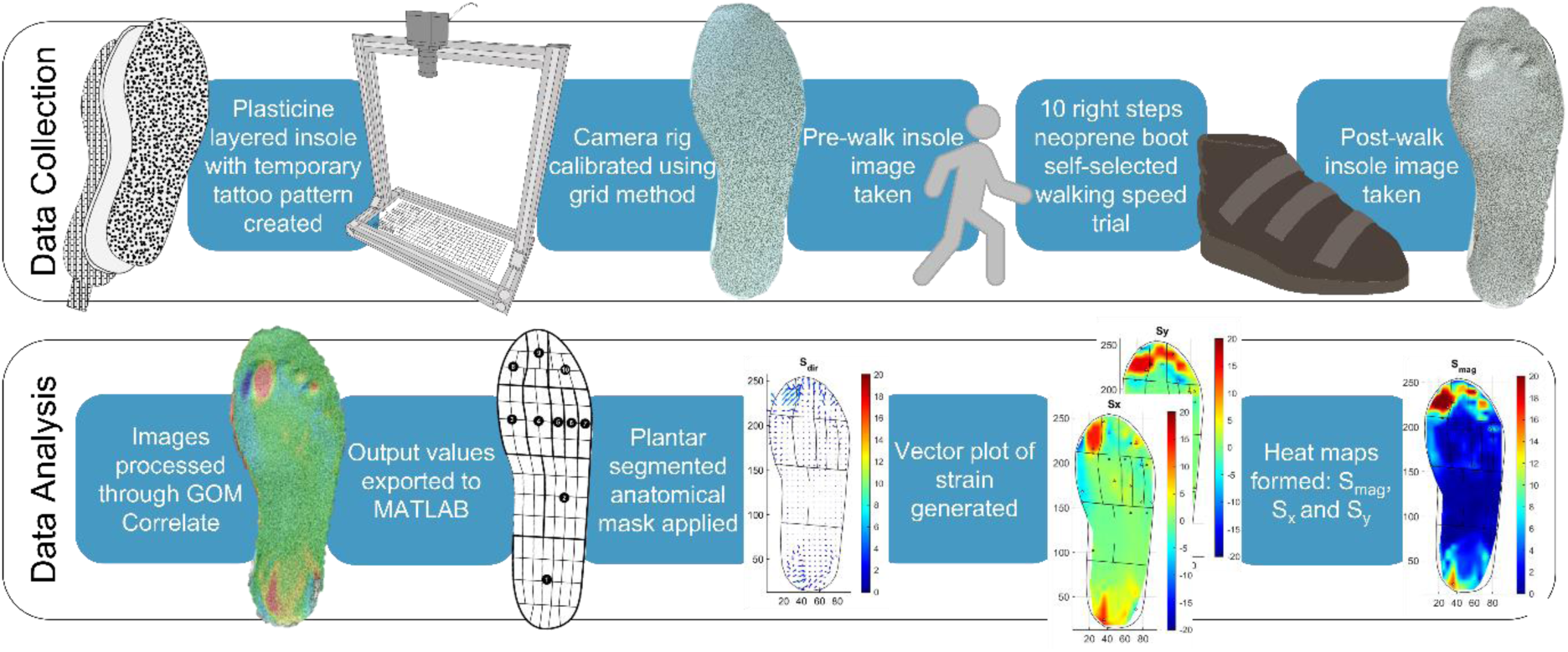
A schematic showing the process of STAMPS data collection and analysis, from initial manufacture through to generation of strain maps and associated summary metrics.

### Insole fabrication

The insole was developed by first identifying an appropriate material for the deformable mid-layer. Preliminary experimentation found that industrial plasticine, an oil-mixed clay composite, provides the requisite mechanical behaviour of plastically deforming under the expected plantar loads found at the foot-surface interface, exhibiting minimal creep and amenable to being formed into regulated laminar sheets which can be cut to a desired size. It has been used extensively to model plastic deformation of metals and flow behaviour during rolling and extrusion due to these properties.^25^ Chijiwa et al., demonstrated that temperature affects the yield stress of plasticine and that it undergoes a period of hardening during the first 24 hours after moulding. After 24 hours, time had minimal effect on the stress-strain relationship.^26^ Accordingly, STAMPS insoles were made at least 24 hours prior to use, and maintained at a constant temperature of 15 degrees prior to use. A commercial clay-roller (CT-500, North Star Polaris) was used to obtain a controlled 5mm thick sheet for fabrication of the complete insole. This rolling process reshaped the bulk plasticine block into laminar form by rolling from an initial ‘thick’ setting (nominal 10mm) towards a target thickness (5mm) in five stages, progressively reducing thickness in each. A cross-patterned nylon mesh (pitch 2mm) was selected as the insole backing to provide a low-profile strain limiting layer. This was affixed to the plasticine layer by including it in the final stage of the rolling process. The mesh extends posteriorly in a tab to prevent adherence to the footwear and allow removal without further deformation, from footwear (see Figure 1).

The speckle-patterned film layer needs to provide a high contrast image while remaining stable over repeated cycles of loading. Our solution was informed by Quino et al., who describe techniques for applying DIC speckle patterns for challenging scenarios.^27^ Accordingly, we printed a speckle pattern onto ‘temporary tattoo’ film (MEDIA-TATTOO-3T, Silhouette America, Inc.) with a thickness of 180um using a commercial inkjet printer. The tattoo film is then applied to the upper surface of the laminar plasticine mid-layer, achieving strong adherence through the thin water-activated adhesive backing. This approach enables repeatable use of standardised high-resolution speckle patterns while having negligible influence on the overall deformation mechanics of the insole.

As a final stage, a standardised sized template and scalpel is used to precisely cut insoles from a laminar sheet without disruption of the pattern or deformable layer. A dusting of talcum powder is applied to the patterned surface after the adhesive has dried and before storage to mitigate against unwanted adherence. A fine layer is applied that does not interfere with DIC tracking or strain analysis.

### DIC Methodology

The basis of effective DIC is an appropriately patterned sample coupled with an imaging system which can capture sufficiently detailed information for subsequent processing.^18^ To meet these demands, a custom digital-camera platform was built to provide a repeatable means of imaging insoles of varying size, as shown in Figure 1. The camera-platform is based on a USB camera (Ultra HD IMX317 USB Camera, ELP Ltd.) using a high-quality charge coupled device and fixed focus lens to obtain 4K (3840×2160) images. The camera is fixed perpendicular to the imaging surface at a height to ensure the desired field of view is captured entirely. The MATLAB camera toolbox was used to calibrate the system by identifying the intrinsic camera parameters in this configuration. The same toolbox was then used in a custom script to capture and return undistorted images of insoles for subsequent DIC analysis.^28^

Stochastic speckle patterns help to standardise the DIC process across multiple insoles, avoiding the variability inherent in manual ‘spray patterning’ techniques. Patterns are defined in terms of parameters of *speckle size, speckle density and variability*. The DIC process then determines strain based on tracking *‘subsets*’ of the patterned region of interest (ROI) which must contain at least 3 speckles. Subsets are distributed across the ROI according to the DICs ‘*step size’* which allows some overlap between subsets and effectively defines the resolution of the resultant strain data.^29^ For this application these pattern and DIC parameters were selected based on recommendations for best practice in the literature.^30 29^ Thus, we used a commercial pattern generator (Correlated Solutions Inc.) to generate a speckle pattern with 65% speckle density, speckle size of 0.8mm and pattern variation of 75%. The associated DIC parameters were a 40 pixel subset and 12 pixel step-size. With the imaging configuration described above, this provides speckles of 9 pixels (fitting for robust tracking) with an average of 15 speckles per subset. This provides high resolution and attenuates noise without a prohibitive computation time.^29^ The DIC process was implemented using commercially available DIC software (GOM correlate, 2020). Pre- and post-images of each insole were imported into the software and used to generate a strain map using the above parameters. The resultant pointwise strain data were then exported into a CSV data file which defined εx and εy for each node of a 1 mm spaced grid for subsequent segmentation and post processing.

### Segmentation and post processing

A custom analysis script (MATLAB, Mathworks) was developed to post-process the computed strain data, providing opportunity for clinically-specific analysis and visualisation.

Firstly, the plantar strain data are pre-processed to remove outliers caused by artefacts in the physical measurement process (e.g. due to disruption of the insole edges) or in the DIC analysis.

The strain map is then segmented by dividing the plantar aspect into 10 regions according to a mask definition employed in commercial plantar analysis software (PEDAR INC): Heel, midfoot, 1^st^ Metatarsal Head (MTH), 2^nd^ MTH, 3^rd^ MTH, 4^th^ MTH, 5^th^ MTH, hallux, second toe, toes 3-5.^31^ The mask is scaled and rotated to fit the specific plantar data (i.e. to accommodate insole size variance) after which the strain data are allocated into each region. The segmented data are aligned with standard anatomical axes (medio-lateral in X and antero-posterior in Y), as shown in Figure 1.

After segmentation, summary metrics are calculated for each region, and the overall plantar space, to identify peak strains in the medio-lateral axis (min S_ML_, max S_ML_), antero-posterior axis (min S_AP_, max S_AP_) and strain magnitude (max S_MAG_). These summary data were exported for statistical analysis during each study. In conjunction, the segmented strain data were used to generate colour-mapped surface plots of S_ML_, S_AP_, S_MAG_ showing peak locations (see Figure 1).

## Experimental Evaluation of STAMPS

To investigate the efficacy of using STAMPS as a measurement tool, and to characterise its operating behaviour, experimental testing was conducted. The aim of the testing was to evaluate the effects of environmental conditions (temperature), operating conditions (varying load regimes of pressure, shear stress, load cycles) and insole thickness. The outcomes from this testing were then used to inform the methods employed in the pilot study reported in *Pilot Study of STAMPS*.

### Methods

An experimental test process was developed by using a Universal Mechanical Tester (UMT) system (UMT TriboLab, Bruker) to apply controlled load regimes to insole samples, as shown in Figure 2. The UMT comprises a load-controlled indenter (Z axis) and a reciprocating plate (X axis) driven by position-controlled micro-positioning stages. The UMT measures load in each axis at 100 Hz during movement. Insole samples were fabricated as described above and held on the reciprocating plate using a custom 3D printed fixture. The indenter was fitted with a 3D printed hemi-ellipsoidal cap (35mm diameter) to approximate the dimensions of the 1^st^ MTH and overlying soft tissue. Scripts were written for the UMT to define the desired load regimes. The applied load regimes were selected based on the anticipated loads at the foot-surface interface. This was informed by Brady et al. who performed compressive testing of foot specimens with loads between 20 and 30 N.^32^ The test setup was maintained at room temperature (22 °C) throughout the process.

**Figure 2:**
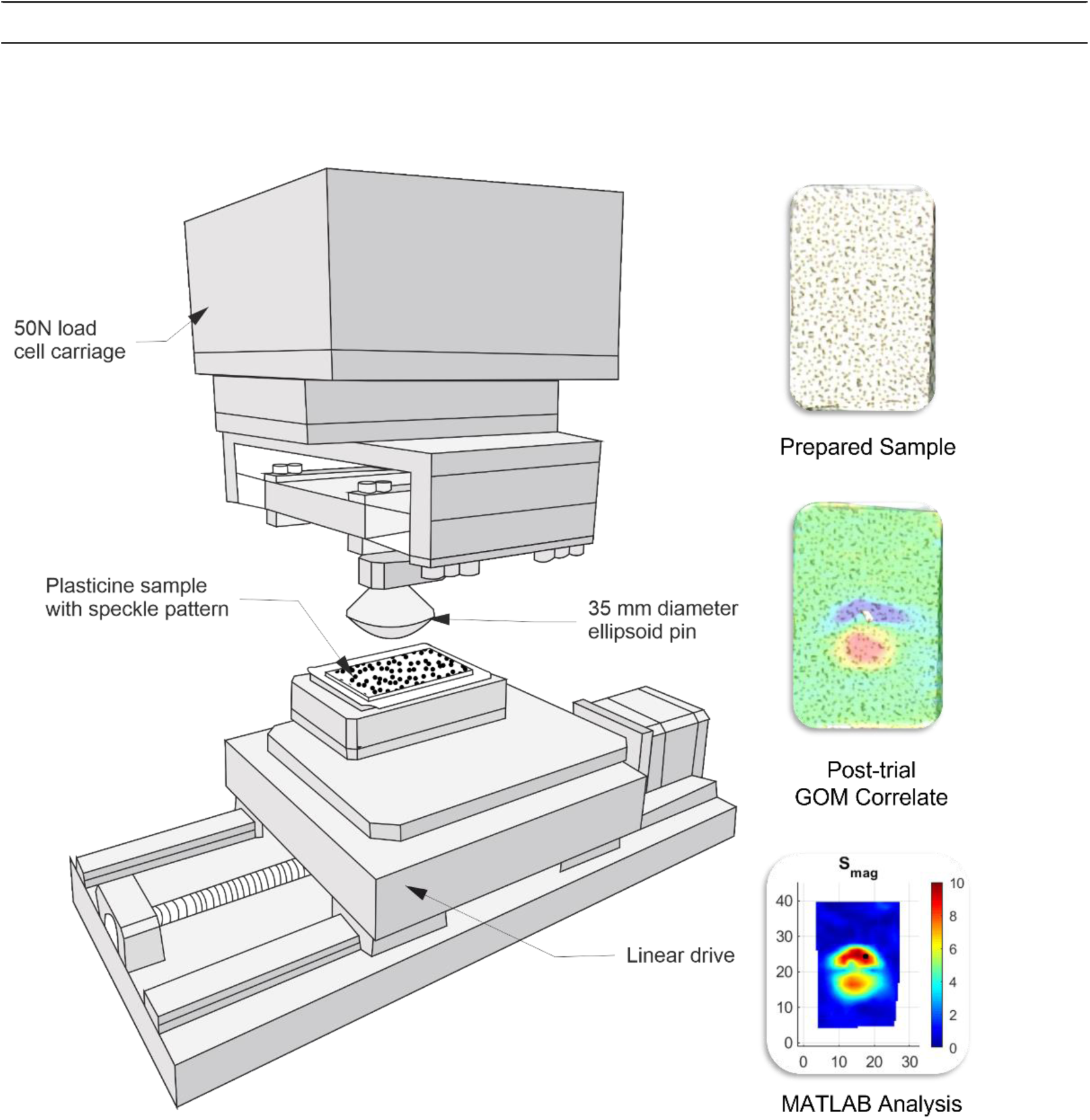
A schematic depicting the UMT (Tribolab, Bruker) configuration used in the experimental evaluation studies and showing an example of the data processing pathway of a single sample.

An experimental matrix, summarised in Table 1, was developed to investigate the various conditions’ experimental parameters around a baseline configuration. Three repeats were conducted for each configuration. Normal and shear loading were defined as a displacement based on preliminary testing to produce an equivalent range of strains to those observed with in-shoe testing. Images were taken of each sample pre and post each loading regime (as defined in Table 1), for subsequent DIC analysis.

**Table 1:** The testing matrix of experimental conditions used to conduct UMT studies for the experimental evaluation of STAMPS.

| Parameter Varied | Range | Secondary Parameter | Cycles |
| --- | --- | --- | --- |
| Normal Load | [10,20,30] N | Shear Load | 1-10 Cycles |
| Shear Load | [0,0.75,1,1.25] mm | Normal Load |  |
| No. Load Cycles | [1,2,3,4,5,10] cycles |  |  |
| Insole Thickness | [4,5,6] mm |  |  |
| Temperature | [5,10,20] °C |  |  |
| Baseline: Normal Load = 20 N, Shear Load = 1mm, Thickness = 5mm, Room Temperature = 22 °C |  |  |  |

Each sample was analysed using the DIC methodology described in *System Development* to obtain a strain map and associated summary metrics of the strain minima and maxima, as shown in Figure 2. These were then collated and processed with statistical analyses.

### Results

All test configurations were conducted and processed using the DIC methodology successfully. The results are summarised in Table 2 and Figure 3.

**Table 2:** Collated results from UMT evaluation studies showing the effect of force, shear distance, sample thickness, temperature and number of steps on S_MAG_ (%).

| Force (N) | Shear Distance (mm) | Sample Thickness (mm) | Temperature (°C) | Number of Steps |  |  |  |  |  |
| --- | --- | --- | --- | --- | --- | --- | --- | --- | --- |
|  |  |  |  | 1 | 2 | 3 | 4 | 5 | 10 |
| | | | | Effect of Shear Distance on $S_{mag}$ | | | | | |
| 10 | 1 | 5 | 22 | 3.10 | 4.89 | 6.98 | 9.67 | 11.94 | 16.24 |
| 20 | 0 | 5 | 22 | 4.95 |  |  |  | 6.35 | 7.03 |
| 20 | 0.75 | 5 | 22 | 4.66 |  |  |  | 12.54 | 23.44 |
| 20 | 1 | 5 | 22 | 5.52 | 10.07 | 12.85 | 17.44 | 22.08 | 37.27 |
| 20 | 1.25 | 5 | 22 | 6.00 |  |  |  | 21.66 | 32.61 |
| 30 | 1 | 5 | 22 | 7.36 | 9.27 | 14.23 | 20.54 | 24.21 | 40.41 |
| | | | | Effect of Sample Thickness on $S_{mag}$ | | | | | |
| 20 | 0 | 4 | 22 | 5.01 |  |  |  | 6.49 | 7.74 |
| 20 | 0 | 5 | 22 | 4.95 |  |  |  | 6.35 | 7.03 |
| 20 | 0 | 6 | 22 | 6.69 |  |  |  | 8.78 | 10.22 |
| 20 | 1 | 4 | 22 | 5.32 |  |  |  | 18.56 | 31.67 |
| 20 | 1 | 5 | 22 | 5.52 |  |  |  | 22.08 | 37.27 |
| 20 | 1 | 6 | 22 | 5.22 |  |  |  | 19.99 | 28.63 |
| | | | | Effect of Temperature on $S_{mag}$ | | | | | |
| 20 | 0.75 | 5 | 5 | 3.72 |  |  |  | 6.14 | 8.41 |
| 20 | 0.75 | 5 | 10 | 4.30 |  |  |  | 6.81 | 8.37 |
| 20 | 0.75 | 5 | 20 | 4.11 |  |  |  | 5.84 | 11.50 |
| 20 | 1.25 | 5 | 5 | 3.60 |  |  |  | 5.93 | 7.31 |
| 20 | 1.25 | 5 | 10 | 3.15 |  |  |  | 6.79 | 14.62 |
| 20 | 1.25 | 5 | 20 | 4.54 |  |  |  | 13.21 | 21.42 |

**Figure 3:**
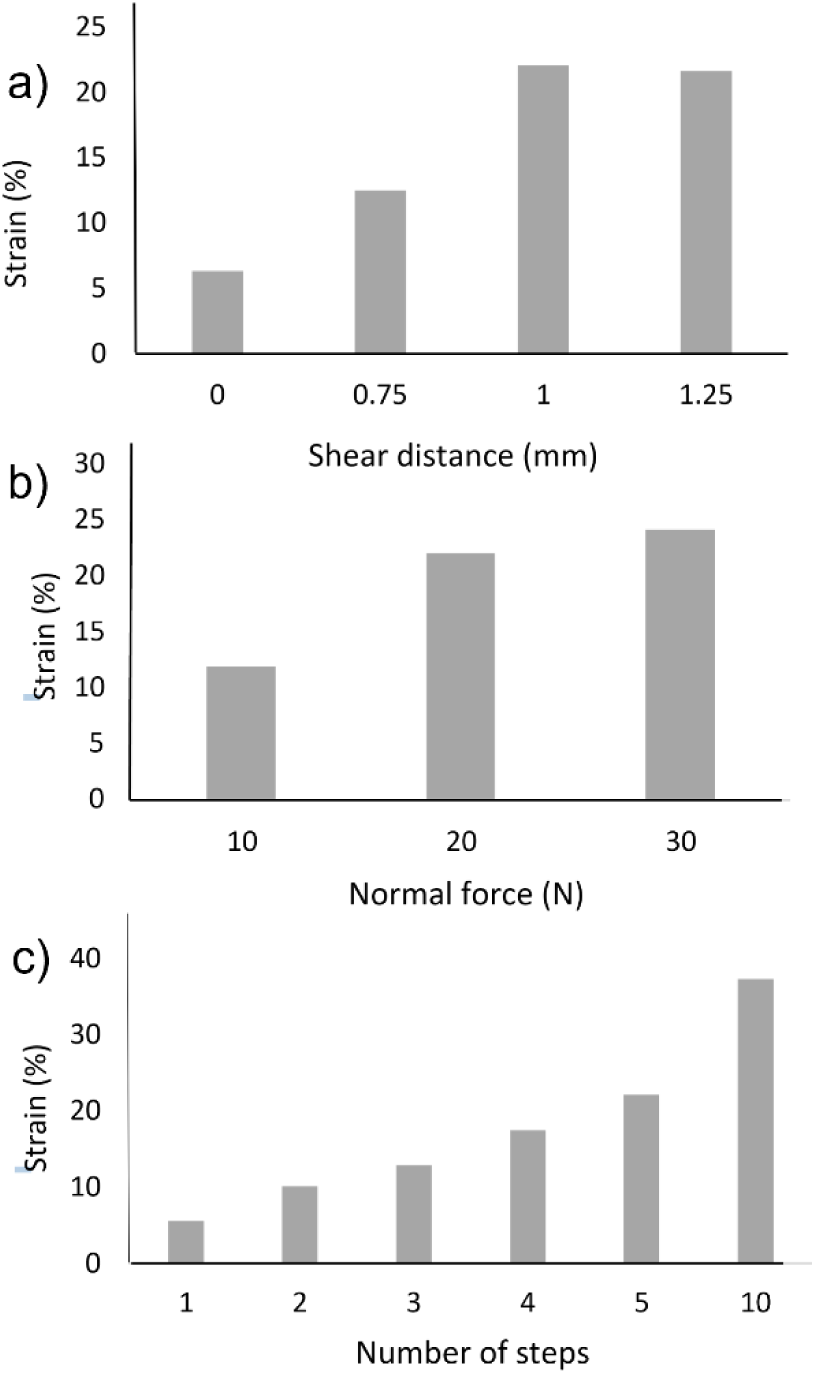
Results of the UMT evaluation studies showing the strain recorded for varying degrees of a) shear distance b) normal force and c) steps applied to the samples.

The results show with increased normal force, at a constant shear distance, S_MAG_ increased in a linear fashion (Figure 3). Furthermore, they showed that with increased shear distance, at a constant force, S_MAG_ increased (Figure 3). The effect was found at up to 1 mm of shear with 20 N of normal force, at 1.25 mm of shear, no further increase was noted. The mean coefficient of variation across tested applied force and strain values with sample thickness 5 mm at a constant temperature was 7.29 %. S_MAG_ was also found to increase in a linear fashion with increased number of cycles.

The results were used to inform a number of parameters for the pilot study. Increased temperature increased S_MAG_ at a constant applied pressure and shear distance; furthermore, lower temperatures (<5 °C) were found to alter the adhesive properties of the temporary tattoo film. Therefore, insoles were subsequently maintained at a constant temperature of 15 °C. Sample thickness had no influence on S_MAG_, this suggests the minimal variation in thickness of insole caused by manually rolling will not affect the results. Outcomes were consistent and pattern integrity maintained to ten steps. The results also validate the technique as a robust method to assess changes in normal force and shear stress.

## Pilot Study of STAMPS

A pilot study was conducted to assess the efficacy of using the STAMPS system to measure in-shoe plantar strain in healthy participants.

### Methods

Ethical approval was obtained from the University of Leeds Ethics committee to conduct the study (LTMECH-005). Eligible participants were age >18 years and capable of walking unaided for 50 metres. Participants were excluded if they had a diagnosis of diabetes mellitus, major or minor lower limb amputation, or significant comorbidities associated with mobility or foot health. Prior to assessment, participants read the participant information sheet and provided written consent. Demographic data, including weight were recorded.

Participant shoe size was measured and the correctly sized supportive neoprene boot (Ninewells Boot, Chaneco inc.) was used. A ‘before’ image was taken of the appropriately sized STAMP insole. This was inserted into the right shoe of the participant. A similarly sized insole was inserted into the left shoe to prevent a discrepancy in insole depth and subsequent alteration of gait. Participants were asked to walk 20 steps along a flat surface, ensuring 10 steps were taken with the right foot at a self-selected, normal walking speed. The insole was removed and an ‘after’ image was taken. This process was repeated three times. Following walking assessments with the STAMP insole, walking assessments were performed using the pedar^®^ (Novel GmbH, Munchen, Germany) in-shoe plantar pressure measurement system. Pressure data was compared with strain data, as measured by the STAMPS system to ascertain validity. No validated in-shoe shear system currently exists and therefore results are not able to be compared with validated in-shoe shear data. Participants were required to walk a distance of 10 m along the same flat surface at their self-selected, normal walking speed.^33^ This process was repeated three times. Outcomes of interest were overall plantar aspect and regional values for S_MAG_, S_ML_, S_AP,_ peak plantar pressure and pressure time integral.

### Statistical Analysis

Data were analysed using SPSS statistical software version 26 (IBM Corp, Chicago, USA). The strain map was segmented into a mask containing ten regions: Heel, midfoot, 1^st^ MTH, 2^nd^ MTH, 3^rd^ MTH, 4^th^ MTH, 5^th^ MTH, hallux, second toe and toes 3-5.^31^ The peak S_MAG_, S_ML_, S_AP,_ for each region of interest and the total plantar surface was extracted for the STAMPS insole. Peak plantar pressure (PPP) and pressure time integral (PTI) for each region and of the total plantar surface were extracted via the multimask application (Novel, GmbH Munchen, Germany), using the previously described mask, for pedar^®^. The Shapiro-Wilk test was used to test for normality of continuous variables.^34^ Pearson’s correlation coefficient or Spearman’s rho was used as appropriate to assess the relationship between S_MAG_, PPP and PTI. A significant relationship was determined if r > 0.4 and p < 0.05. To establish repeatability, the coefficients of variation (CV) of PPP and S_MAG_ were calculated for each region, the mean value of CV was calculated for each subject.^15^

### Results

Designed as a proof concept study three participants were recruited,^35^ all provided informed consent, with characteristics summarised in Table 3. Regional S_MAG_ and PPP data for each trial are shown in Figure 4. The results demonstrate different strain patterns between individuals, which are consistent between trials (Figure 5).

**Table 3:**
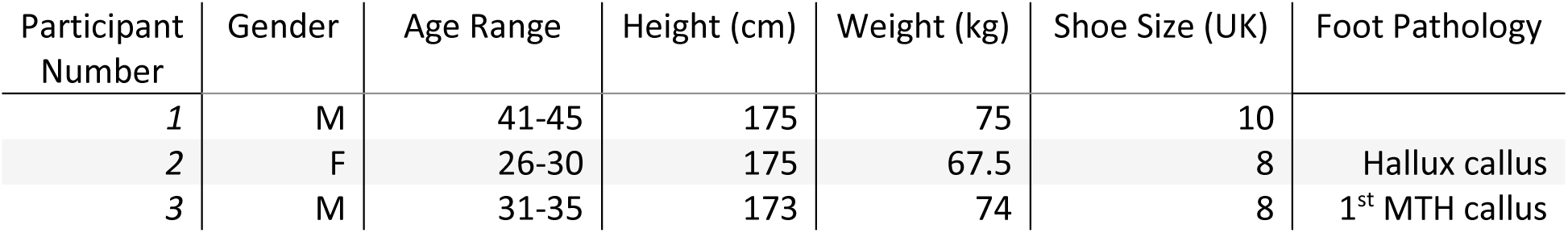
Key characteristics of the pilot study participants

**Figure 4:**
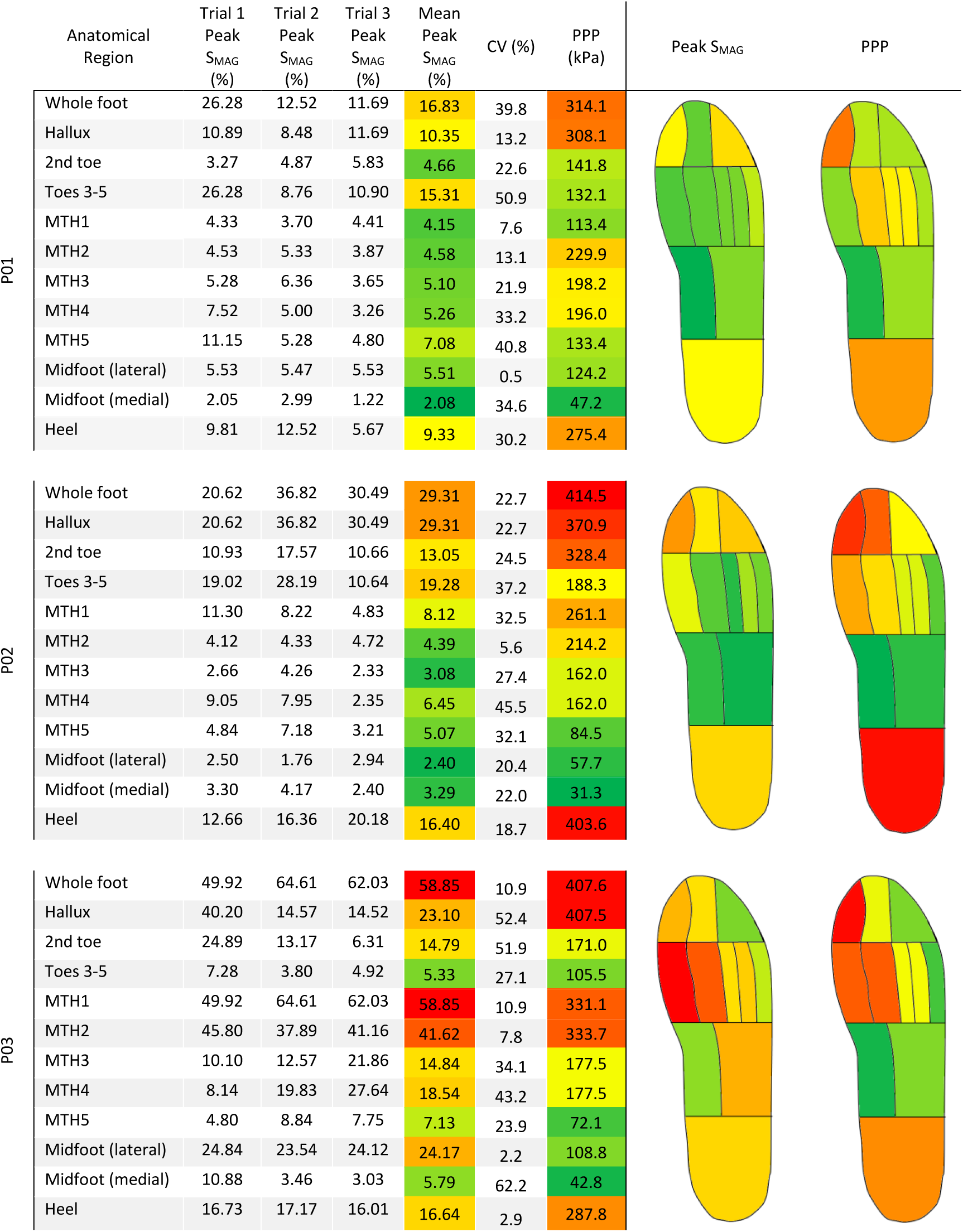
A summary of the plantar loading data showing the regional peak Strain Magnitude (S_MAG_) and Peak Plantar Pressure (PPP) data, together with corresponding graphical representation of the regional data for each pilot study participant (P01-P03).

**Figure 5:**
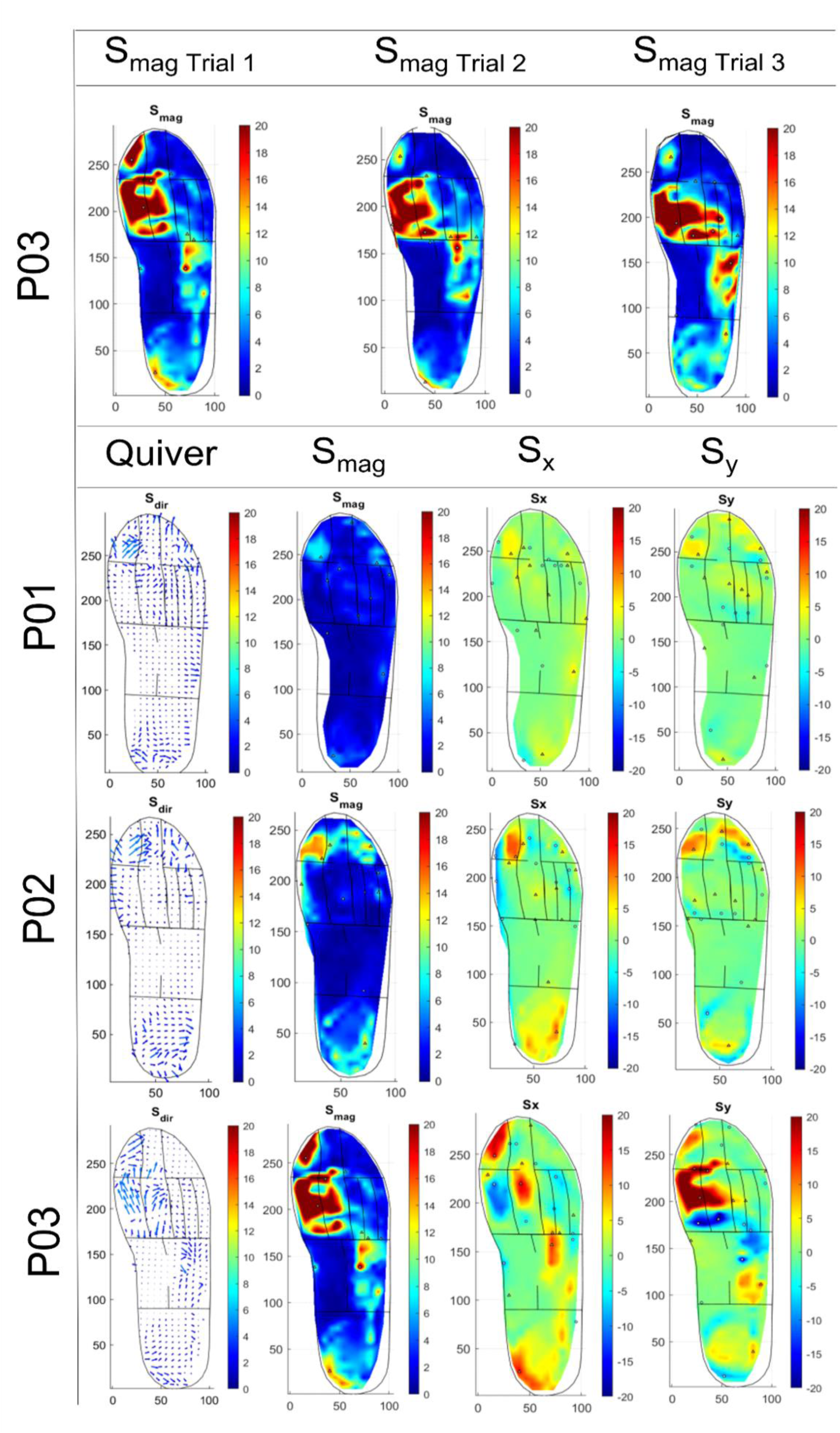
Top: heat maps showing the resultant strain magnitude S_MAG_ for P03 across three repeated trials as a representative example showing consistency of the measurement technique. Bottom: representative examples of strain data for each participant (P01-P03), shown as quiver plots for strain direction (S_x_, S_y_) and heat maps for strain magnitude (S_MAG_) and strain components (S_x_ and S_y_). The XY axes show the insole size in mm, the colour-map scale shows the measured strain values.

Participant 1 demonstrated greatest S_MAG_ in the region toes 3-5 (15.31 %). The area of greatest PPP was the hallux (308.1 kPa). The highest mean S_MAG_ for participant 2 was at the hallux (29.31 %), with high areas of strain also noted at the heel (16.40 %) and region of toes 3-5 (19.28 %). The regions of the hallux and heel were also the regions with greatest PPP (370.9 kPa and 403.6 kPa respectively). Participant 3 showed a different strain pattern, with the highest mean S_MAG_ identified at the 1^st^ and 2^nd^ metatarsal heads (58.85 % and 41.62 % respectively). In contrast to participants 1 and 2, very little strain was found in the region of toes 3-5, and again unlike participants 1 and 2, the region of the heel demonstrated relatively low strain compared with the rest of the plantar surface. The highest PPP was found at the hallux (407.5 kPa), with relatively high pressures at the 1^st^ and 2^nd^ metatarsal heads (331.1 kPa and 333.7 kPa respectively). Again, areas of high strain were found in the absence of high PPP, most notably in the region of the lateral midfoot. Strain patterns were consistent across the three walking assessments for each individual with a mean CV of 26.4 %. The mean CV for PPP was 13.0 %.

The relationship between S_MAG_ and PPP is shown in Figure 6. PPP was normally distributed; however S_MAG_ was not, and therefore Spearman’s rho was used to assess the relationship between variables. Increased PPP was strongly associated with increased S_MAG_; Spearman’s correlation coefficient 0.673 (p <0.0001). Increased PTI was also associated with increased S_MAG_, Spearman’s correlation coefficient 0.653 (p <0.0001).

**Figure 6:**
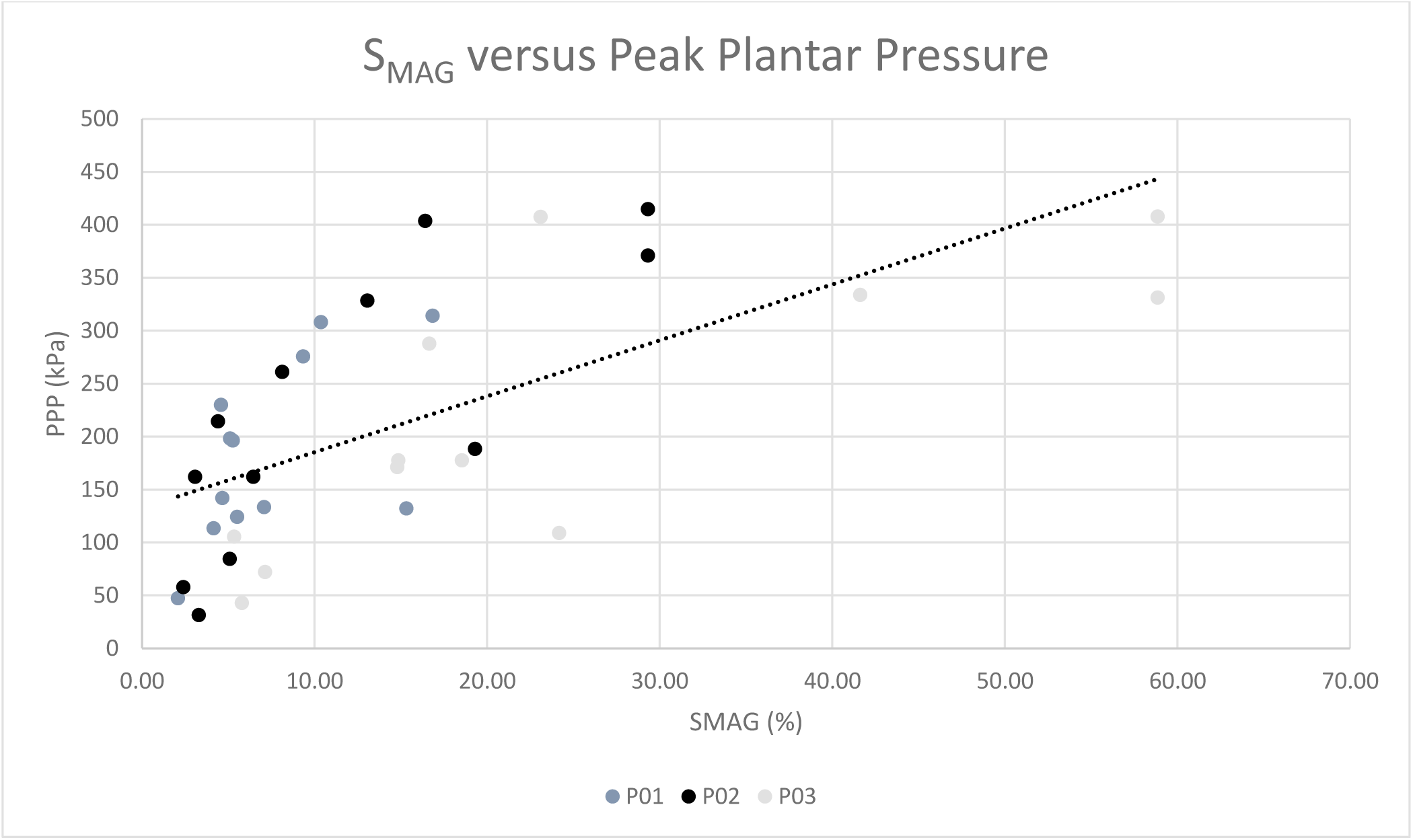
A scatter plot showing the relationship between strain magnitude (S_MAG_) and Peak Plantar Pressure (PPP) for each participant (P01-P03) in the pilot study. The linear trend line shows the reported correlation between these variables with a Spearman’s coefficient 0.673 (p <0.0001).

## Discussion

Development of a low cost and time efficient in-shoe system to measure plantar shear stress and plantar pressure will aid risk assessment and guide treatment strategies for prevention and management of DFUs. As our recent systematic review has shown, few studies describe systems for in-shoe plantar shear stress assessment and no commercial devices have been developed.^13^ Plantar pressure assessment is recommended in the IWGDF guidelines for the prevention of DFUs,^9^ however pressure assessment alone is a poor predictor for DFU development in prospective studies.^6^ Furthermore, there is only a weak association between area of elevated plantar pressure and site of ulceration.^10^ As such there is a requirement for in-shoe systems measuring both plantar pressure and plantar shear stress to determine the contribution of shear stress to DFU development and deterioration. This study reports on a novel technique developed for the measurement of plantar load at the foot-surface interface. The results of the UMT tests support the hypothesis that the strain of the plasticine insole, measured using DIC, occurs as result of normal pressure and tangential shear stress. Exposure to increased levels of normal force and increased shear distance resulted in elevated levels of strain. Strain plateaued above a shear distance of >1 mm due to the loading profile of UMT, which results in point loading and translation. This creates adverse distortion of the pattern, not seen in loading during gait. Consequently, this defined the UMT test regime. Furthermore, it is shown to be repeatable, with a coefficient of variation of 7.29 %.

The method described contrasts with the trend in plantar sensing technology. Strain-gauge systems,^11,14,36–38^ piezoelectric transducers,^39^ magnetic resistive transducers,^16^ optical methods and microstrip antennas all rely upon sophisticated yet costly sensing technology to provide real-time shear stress data. The STAMPS system provides an alternative approach to plantar load assessment, which is low cost, simple and quick to implement. While it does not provide real-time (instantaneous) plantar pressure or shear data, it does record the cumulative strain following a period of gait, from which inferences regarding plantar load can be made. The insoles performed well under test conditions; pattern integrity was maintained throughout with no loss of DIC tracking. Insertion and removal of insoles from test shoes was achieved with minimal difficulty and facilitated by the scaffolding layer. Participants reported no disturbance to gait whilst wearing the insoles. The described method uses two programmes for data processing and analysis; DIC performed using GOM Correlate prior to export to MATLAB for segmentation and peak strain analysis. GOM is a free, easy to use software which generates a heat map that is simple to interpret. In a time-constrained clinical setting, this data may be sufficient for decision making. If regional strain data is required, secondary analysis using MATLAB can be performed. The process can be streamlined, performing both DIC and analysis using MATLAB, however the user-friendly functionality and output of GOM enables third party use with little additional expertise.

The experimental protocol was finalised following extensive preliminary testing and was designed to ensure consistent data collection. Evidence from Chijiwa et al., supported the need to manufacture insoles >24 hours prior to use.^26^ The effect of change in temperature upon resultant strain was demonstrated during UMT testing; as such, insoles were maintained at a constant controlled temperature prior to use. The strain metrics measured by STAMPS are the cumulative effect of a period of gait. Participants therefore must take the same number of steps for comparisons to be drawn. A period of mid gait steps are required, between initiation and termination of gait. Increasing the number of steps increases the proportion of deformation that represents ‘normal gait’, as initiation and termination steps are highly variable.^40^ Preliminary work demonstrated that increasing the step count beyond this limit (i.e. 15-20 steps) degraded the pattern beyond the limit to which it could be reliably tracked, particularly in areas of high plantar strain. It is hypothesised that peak strain will be higher in patients with abnormal foot biomechanics than healthy participants. Therefore, to support future clinical use the limit of strain should not be reached within use in healthy participants. No regions of dropout occurred when healthy participants completed a total of 20 steps, with 10 steps on the ipsilateral (measured) side. Eighty percent of the steps taken during a 20 step walking assessment constitute mid-gait steps, therefore this was preferred over a 10 step walking assessment. For in-shoe plantar pressure assessment, Kernozek demonstrated that eight steps were required to achieve high reliability (>0.90 reliability).^41^ Therefore 20 steps, ensuring 10 on the ipsilateral (STAMPS insole) side was selected.

Patterns of strain distribution and magnitude were consistent for each participant and demonstrated significant variation between individuals. Peak S_MAG_ varied considerably between individuals, with the level of peak S_MAG_ three times greater with participant 3 compared with participant 1. Participant 1 sustained highest peak strain at the regions of toes 3-5, hallux and heel with low levels of strain throughout the remaining regions. Participant 2 sustained highest peak strain at the hallux, with high strain also noted at the regions of toes 3-5 and the heel. In contrast, participant 3 sustained peak strain at the 1^st^ MTH, with elevated levels of strain at the medial metatarsals, with strain reducing moving laterally. Unlike participants 1 and 2, participant 3 also sustained relatively high levels of strain in the midfoot. Direction of strain also varied. Participant 3 sustained considerable, anteriorly directed Sy in the anterior aspect of the 1^st^ and 2 MTH regions, yet S_MAG_ was directed posteriorly at the posterior aspects across the metatarsal head regions, with low values of S_ML_. Participant 2 sustained elevated levels of medially and laterally directed strain at the hallux, yet at the toes, the strain was largely directed anteriorly. Consideration of the patient and patient’s foot pathology is required to interpret plantar load. Further work is required to determine the relationship between the plantar load measured using STAMPS and foot pathology.

A strong correlation was noted between resultant strain and peak plantar pressure, reinforcing this technique as a valid measure of plantar load. Several regions experienced disproportionately greater strain when compared with peak plantar pressure; these included the region of toes 3-5 for participant 1 and 2 and the regions of the 1^st^ and 2^nd^ metatarsals and the lateral midfoot of participant 3. It is hypothesised that the increased strain within these regions is likely as a result of the effects of shear stress, however due to the lack of a valid in-shoe shear stress device, this could not be confirmed. There is also a lack of evidence within the literature to support or refute this hypothesis as no studies have performed pressure and shear stress assessment throughout the entirety of the plantar surface. A cross sectional study of healthy individuals is required to determine the ‘normal values’ of strain and assess the influence of confounding variables including, but not limited to gait speed and weight. Following this, the technique will be used to assess plantar load in an at-risk diabetic cohort.

### Limitations

There are some limitations associated with the STAMPS technique. The strain metrics measured reflect the deformation of the insole following a period of gait. The strong correlation with peak plantar pressure and the results of experimental validation demonstrate that the magnitude of strain is the product of plantar pressure and shear stress at the foot-surface interface. However, direct measurements of peak plantar pressure and shear stress are not performed, and unlike traditional sensing techniques, real-time (instantaneous) data are not recorded.

Variation in strain response was identified during UMT testing with constant application of normal force and shear stress. During in-shoe tests, this variation increased to a mean of 26.4%. Variation within a simulated environment, with controlled application of load was 7.3%. This suggests the high CV is as a result of true variation, rather than error in measurement. Furthermore, the greatest variation was noted at the lower measures of load, making the variation less clinically relevant. The technique uses DIC, which can be subject to systematic errors. Several steps have been taken to ensure that the differences in strain patterns observed are a result of true variation rather than systematic errors. Patterning is consistent, with high contrast between speckle and background. The speckle size, density, variation, facet size and subset spacing has been optimised to reduce the likelihood of error. Furthermore, variables affecting the material properties of the insole including temperature and ‘cure time’ were controlled. As described, further evaluation with a greater number of participants will add further insights into the repeatability of the technique.

This study was designed as a pilot study to demonstrate the principle of the insole system. It was not designed to investigate the effects of covariates that may affect resultant strain metrics. A healthy participant study, involving a larger sample size will be performed to establish normal parameters, and investigate the relationship between strain, weight and walking speed.

The experimental work found increased storage temperature resulted in increased levels of strain recorded with constant shear and normal force applied. Due to the limited period of time the insole is in contact with a participant’s foot, variability in foot temperature is not expected to significantly influence strain outcomes. However, it does have implications for use within a clinical environment, as it requires maintenance of insoles at a constant temperature immediately prior to use. Future development will investigate materials which are less sensitive to temperature as well as the effect of walking speed and weight on resultant strain.

## Conclusions

This study has demonstrated the efficacy of a novel method to assess plantar load across the plantar surface of the foot. Experimental testing validated the method to measure both vertical pressure and tangential shear stress. This technique was successfully incorporated into the STAMPS insole to reliably measure and quantify the cumulative degree of strain sustained by a deformable insole during a period of gait, which can be used to infer plantar loading patterns. Further work is required to establish ‘normal values’ within a healthy population before investigating strain patterns and parameters in patients with diabetes at risk of developing DFUs.

## Data Availability

All data produced in the present study are available upon reasonable request to the authors.

## Acknowledgements

The authors would like to thank the NIHR MIC in Surgical Technologies for their support and Steeper Ltd for provision of custom orthotic shoes used in the testing.

## Ethical approval and participant consent

Ethics were obtained via the University of Leeds Engineering and Physical Sciences joint Faculty Research Ethics Committee. Ethics Reference: LTMECH-005. Consent was obtained from participants of the pilot study.

